# Elastokines are Associated with a Poor Prognosis in Idiopathic Pulmonary Fibrosis

**DOI:** 10.1101/2025.06.06.25327886

**Authors:** DJ Nagel, Takuma Okutani, Samia Lopa, TJ Mariani, PJ Sime, RM Kottmann, Denise Hocking

## Abstract

**Background:** During the development of idiopathic pulmonary fibrosis (IPF), elastin is broken down and replaced with a stiffer substrate which interferes with normal breathing. This remodeling process releases the amino acids desmosine and isodesmosine and elastin degradation products (EDP), collectively known as elastokines, into the circulation and may act as a surrogate for the degree of matrix turnover. We examined the association between circulating and urinary elastokine concentrations, lung function, and transplant-free survival.

**Research question:** What are the associations between circulating and urinary elastokine concentrations (a marker of mature elastin turnover) with disease progression, as measured by transplant-free survival, in IPF?

**Study Design and Methods:** Concentrations of desmosine and isodesmosine (elastokines) were measured via enzyme linked immunosorbent assay (ELISA) in the urine and serum of healthy volunteers and those with IPF. Samples were obtained from the University of Rochester Medical Center Interstitial Lung Disease (ILD) registry/biorepository (n = 81 with IPF and 24 healthy volunteers). We used linear and logistic regression modeling to determine associations between changes in urine EDP concentrations and lung function. Secondary analyses included the effect of anti-fibrotic medications on EDP concentration and assessment of associations between EDP concentrations at the time of diagnosis, lung function, and clinical outcomes.

**Results:** Patients with IPF were older, more likely to be male, had ever smoked, and had worse lung function compared to healthy volunteers (p value <0.02 for all parameters). Patients with IPF had higher concentrations of elasotkines (p <0.001), and among those with IPF, higher elastokine concentrations were associated with reduced forced vital capacity (FVC, p = 0.026), and decreased three-year transplant-free survival (p=0.0005).

**Interpretation:** Circulating and urinary elastokines are biomarkers of matrix turnover and are associated with relevant clinical outcomes in patients with IPF.

## Introduction

Idiopathic pulmonary fibrosis (IPF) is a disease of unknown cause marked by progressive scarring of the lung occurring almost exclusively occurs in older adults. As the world’s population ages, its incidence and prevalence are on the rise(1). The mechanisms by which IPF develops remain incompletely understood, but it is thought that an injury occurs to airway epithelial cells, leading to fibroblast activation/differentiation, and lung scarring. IPF carries a prognosis only slightly better than lung cancer(2). Currently, there are two FDA-approved medications for the treatment of IPF: nintedanib (a triple kinase receptor inhibitor)(3) and pirfenidone (a partial TGF-β antagnoist)(4). While these medications decrease the rate of decline in FVC and reduce respiratory related hospitalizations, they do not reverse established fibrosis and are often not well-tolerated. The only cure for IPF is lung transplantation(5). However, lung transplant survival rates are the lowest among solid organ transplants; the median survival rate after lung transplantation is 5.8 years(6).

Elastin is a hydrophobic polymer that provides tissue resilience, elasticity, and stability that is essential for proper lung function. During the development of IPF, pro-fibrotic cytokines, like transforming growth factor-beta (TGF-β), induce the activity of elastases and proteinases, which degrade mature elastin, and are increased in the lung tissue of patients with IPF(7). Elastin is synthesized as tropoelastin by fibroblasts, smooth muscle cells, endothelial cells, and chondrocytes(8) and is subsequently condensed to mature elastin in the extracellular space. Mature elastin is a polymer of cross-linked tropoelastin fibrils made of glycoproteins such as latent transforming growth factor β-binding proteins (LTBPs), fibrillin-1 and -2, and microfibrillar-associated proteins (MFAPs)(9). In healthy adult tissues, ultra-long proteins, like elastin, typically have a very low turnover rate (half-life of approximately 74 years)(10). In contrast, most other proteins have half-lives of minutes, hours, or days(11). However, when homeostatic conditions are altered by chronic illness, elastin turnover occurs much more rapidly. Adult tissues generally lack the ability to regenerate functional elastin fibers to counteract this increased turnover(12), which can lead to irreversible changes in tissue structure and function. In addition, although elastin mRNA is stabilized by TGF-β(13), over-expression of tropoelastin via transduction was not sufficient to induce elastic fiber formation(14), indicating that degraded elastin is not easily replaced.

When mature elastin is degraded, its amino acid constituents (isodesmosine and desmosine) and elastin sub-fragments (elastin degradation products or EDPs) (collectively known as elastokines) become liberated, where they can now be detected in body fluids, including urine, plasma, and bronchoalveolar lavage fluid(15). Elastin-related pathology has significant prognostic information as several reports have shown an association between higher concentrations of circulating EDPs and increased mortality(16, 17). Indeed, increased elastin remodeling has been reported in diseases such as chronic obstructive pulmonary disease (COPD)(18, 19), pulmonary arterial hypertension(20), abdominal aortic aneurysm, and rheumatoid arthritis(21). It has also been shown that chronic diseases accelerate elastin turnover relative to age-matched controls(22).

Here we measured elastokine concentrations in patients with IPF compared to healthy controls and studied their association with relevant clinical outcomes.

## Study Design and Methods

Study sample: The University of Rochester Medical Center ILD registry/biorepository was created in 2008. The diagnosis of IPF was designated either according to the current professional guidelines or through multi-disciplinary discussion(34-38). Patients who expressed an interest in participating were consented at any time in the process of their disease and could donate biospecimens serially. For this analysis we used the first donation that was either the closest to when consent was obtained or when the diagnosis was first documented (often occurring at the same visit).

Clinical outcomes (lung transplant or death) were then observed for a period of three years after initial sample collection. If the patient survived or did not receive a transplant during those three years, lung transplant-free survival was documented.

Urinary and circulating elastokine measurement: The concentration of amino acids desmosine and isodesmosine was measured primarily in urine and serum via a commercially available enzyme-linked immunosorbent assay (ELISA) kit (mybiosource.com, Catalog # MBS702024 and MBS702214, respectively). Urine samples were available for the entire cohort (n=81 patients with IPF, n=24 healthy controls), and serum samples were available from a subset (n=24 patients with IPF, n=8 from healthy controls). We chose to use urine as the primary source because it is the least invasive and most widely available biospecimen in the repository. Samples were stored at −80° C until they were thawed for use. Duplicates for standards and each patient sample were used, and elastokine measurement was performed according to manufacturer instructions using replicates for each donor. The product literature states that for both assays, the intra-assay coefficient of variation is < 8% and the inter-assay precision has a coefficient of variation of < 10%. Quatitative measurements were then obtained based on absorption at 450 nm with an iMark microplate reader (Bio-Rad). Results were then analyzed and graphed using GraphPad Prism (v10) or SAS (v9.4).

Physiologic Assessments: Spirometry (forced vital capacity, FVC) and diffusing capacity for carbon monoxide (D_L_CO) were performed as part of standard clinical care, in accordance with current professional guidelines(39-41). Patients typically underwent lung function testing on the day of their clinical appointment; however consent, enrollment, and sample donation may have occurred on a different day from their clinic appointment. In these cases, spirometry values that were obtained closest to the enrollment date were considered the baseline values for each participant.

### Adjudication of Death and Lung Transplantation

Death and lung transplantation status were adjudicated by the study personnel (investigators and study coordination staff). Time to censored event occurred for a period of 3 years following the initial date of sample donation.

### Statistical Analysis

Based on a prior study demonstrating the effect size of fold change in EDP concentration to be 10 units per 1 standard deviation(8) and confirmation of this effect size from our preliminary data, a patient sample size of 63 was deemed necessary to detect the same effect size at 80% power and a two-sided 0.05 significance level. Socio-demographic and clinical variables investigated include FVC, D_L_CO, smoking status, gender, ethnicity, race, age, presence of hypoxemia, and anti-fibrotic medication use. Linear and logistic regression modeling was utilized to determine associations between changes in urine EDP concentrations and lung function. Secondary analyses included the effect of anti-fibrotic medications on EDP concentration and assessment of associations between EDP concentrations at the time of diagnosis and clinical outcomes (time to respiratory-related hospitalization, transplant, or death). For the latter, we used time to event analysis to assess 3-year transplant-free survival. To create the comparator groups, we divided elastokine concentrations into tertiles. The GAP (Gender, Age, and lung Physiology) staging system was initially created(42) and subsequently validated(43) to assess 1, 2, and 3-year mortality in patients with IPF. We assessed EDP concentration as a function of GAP stage to provide additional support for the prognostic implications of elevated EDP concentrations.

It is unknown if there is a relative difference in amino acid release between desmosine and isodesomine under conditions of elastinolysis. To assess the precision of urinary isodesmosine versus desmosine to detect a change in EDP concentrations, we selected a random sub-population and performed a correlation analysis based on urinary ELISA concentrations. It is also unclear if there are relative differences in EDP concentrations based on specimen source. To assess the accuracy of desmosine measurement between urinary and serum sources we used Bland-Altman analysis, using a different set of randomly selected patients.

All statistical analysis was performed either with GraphPad Prism (v10, DJN) or SAS (v9.4, SL) software packages. Statistical significance was considered when p < 0.05. Most figures were created using a combination of the above programs and BioRender (biorender.com)

## Results

*Baseline Demographics:* Compared to healthy subjects, patients with IPF were older (71 vs 40 years of age, p <0.001)), more likely to be male (71.6% vs 37.5%, p =0.004), more likely to have ever smoked (69.1% vs 41.7%, p =0.021), and have a higher BMI (28.0 vs 24.3 kg/m^2^, p =0.003). Patients with IPF were more likely to be male (71.6% vs 28.4%, p =0.0001), Caucasian (97.5% vs 2.5%, p <0.0001), and have ever smoked (69.1% vs 30.9%, p =0.0006). Compared with healthy volunteers, patients with IPF had worse lung function (FVC 72% vs 99% predicted, p <0.001; DLCO 46% vs 82% predicted, p=0.08), a higher supplemental oxygen need (53.1% vs 0%), higher mortality (65.4% vs 0%) and more frequent transplant rates (12.35% vs 0%). Approximately 46% of patients with IPF were taking an anti-fibrotic medication.

### Association of Urinary Elastokines and IPF

To assess for differences in concentrations of desmosine and isodesmosine, we first analyzed urine samples from 81 patients with IPF and 24 healthy volunteers for desmosine concentrations. The average urinary desmosine level in patients with IPF (1.4 ng/mL) was significantly higher than in healthy volunteers was (0.321 ng/mL, Figure 1A, p<0.0001).

**Figure 1.**
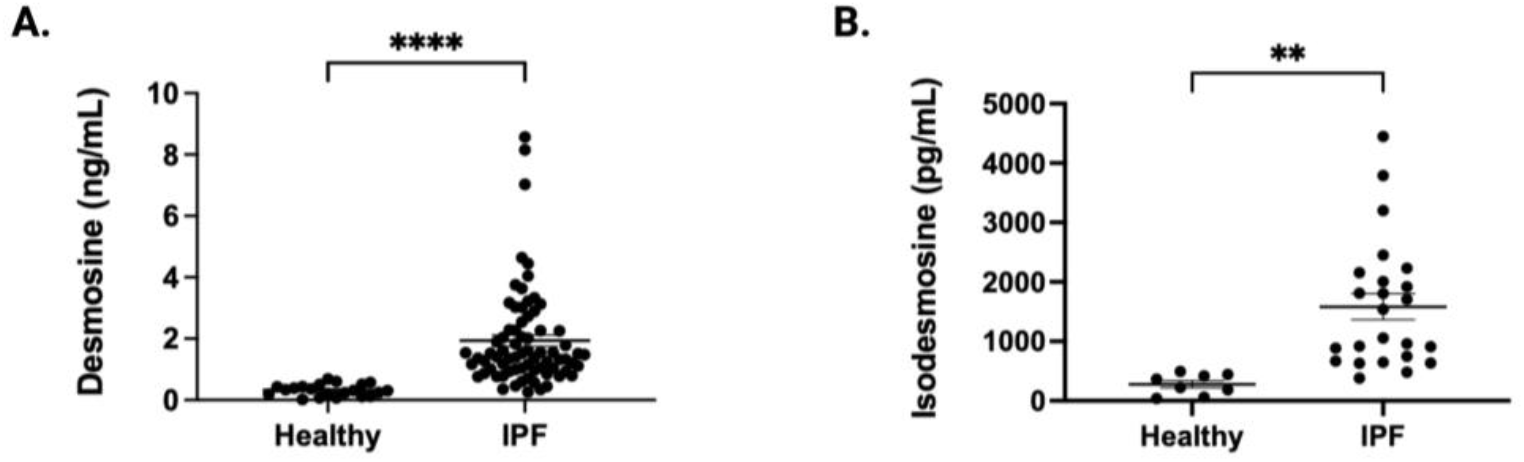
Elastokine concentrations are elevated in IPF. Urinary concentrations of desmosine and isodesmosine were compared among healthy volunteers and patients with IPF. Both desmosine (A) and isodesmosine (B) concentrations are significantly higher in patients with IPF. Desmosine p (****) < 0.0001 and isodesmosine p (**) 0.002.

To assess for any potential difference in urinary elastokine concentrations, we then measured isodesmosine concentrations in a randomly selected subset of healthy individuals and those with IPF. Isodesmosine concentrations were also significantly elevated in patients with IPF (Figure 1B, p=0.002). This data demonstrates that people with IPF have higher urinary concentrations of both desmosine and isodesomine, suggesting they have higher rates of mature elastin fiber turnover.

### Assessment of Accuracy and Precision of Desmosine and Isodesmosine concentrations

It was previously unknown if urinary desmosine and isodesmosine concentrations are similarly elevated in patients with IPF. In theory, if one amino acid is more sensitive at detecting elastin turnover, then this could have potential clinical implications. Figure 1 demonstrates that both are significantly elevated, but we wanted to assess this at a more granular level. Therefore, we analyzed the correlation between these two amino acids between patients with IPF and healthy volunteers. As seen in Figure 2A, there was no significant difference when comparing urinary concentrations in either group. There was moderate correlation between urinary desmosine and isodesmosine in both cohorts (Figure 2B, pearson r coefficient of 0.5123).

**Figure 2.**
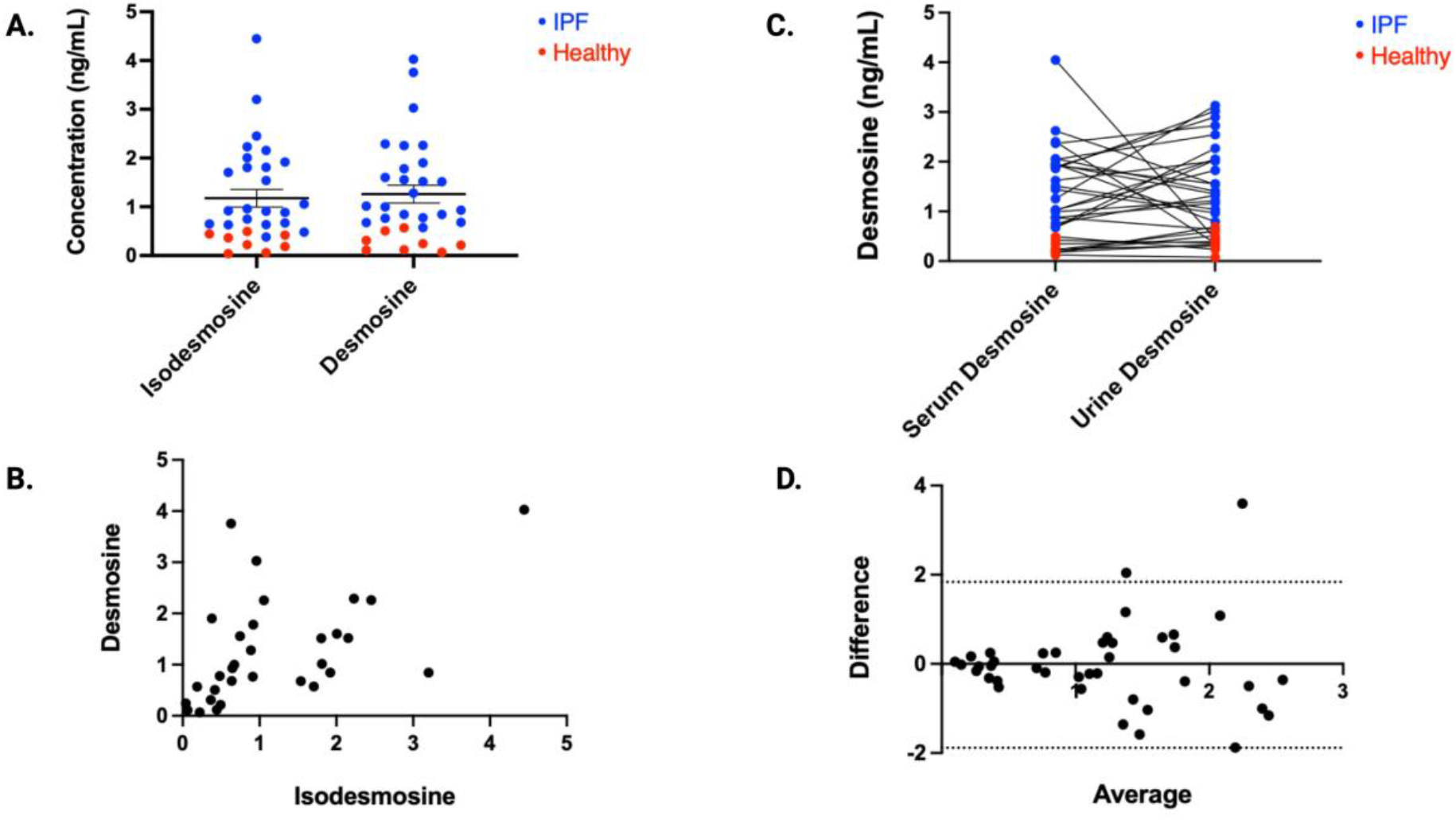
Elastokine concentrations are similar regardless of source. Urinary concentrations of isodesmosine and desmosine were compared (A and B). The relative concentrations of desmosine were compared by source as well (C and D). There was no significant difference between isodesmosine and desmosine measurements (A) and pearson r coefficient was 0.5123 [0.193 – 0.7335] (B). There were no stastical differences in desmosine concentrations when measured from serum or urine (C). Bland-Altman analysis (D) revealed a bias of −0.049 (Limits of Agreement: −2.117 – 2.019).

We next wanted to assess for differences in desmosine by specimen source and used biospecimens from matched patients to compare urinary and serum desmosine concentrations. As expected, concentrations in healthy volunteers were lower whether measured in serum or urine. While concentrations of both were higher in patients with IPF, there was no statistical difference between the two sources from the same donors (Figure 2C). We then compared for possible differences between desmosine and isodesmosine concentrations from the same source (urine) and found good agreement between both elastokines (Figure 2D). Taken together, these data indicate that urinary EDPs are a faithful reflection of serum EDP concentrations in healthy controls and patients with IPF.

### Association of Elastokine Elevation and Lung Function

The change in FVC over time has been used as a primary endpoint in clinical trials assessing the efficacy of medications for the treatment of IPF and has been correlated with mortality(44). Thus, we aimed to investigate the relationship between elastokine elevation and lung function. We found that higher EDP concentrations are associated with worse lung function, as measured by FVC (Figure 3A). While a similar correlation was observed with D_L_CO (Figure 3B), this did not reach statistical significance. Patients with higher concentrations of desmosine were more likely to be taking an anti-fibrotic medication (OR 2.057 [0.666-6.28] vs 0.4861 [0.1591 to 1.50], data not shown), but since some samples were collected prior to anti-fibrotic approval, we did not perform additional analysis and are unable to assess the effect of anti-fibrotic medications on elastokine concentrations over time.

**Figure 3.**
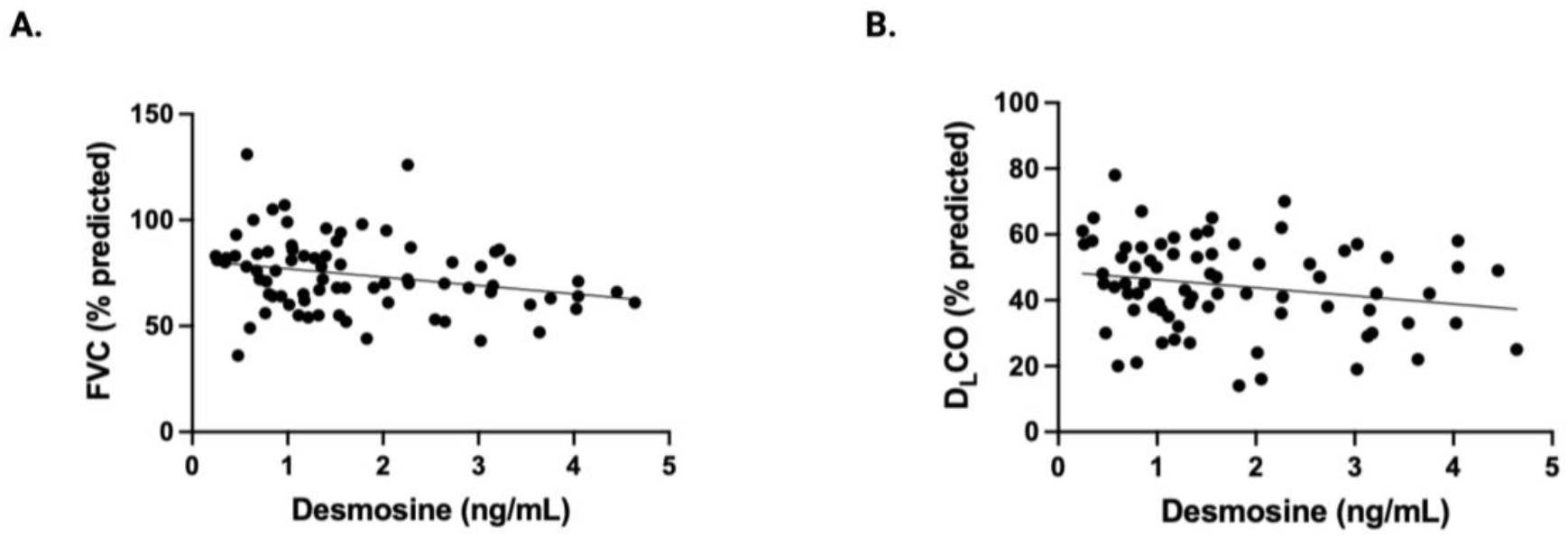
Elastokine concentrations are inversely associated with Forced Vital Capacity. Higher concentrations of desmosine are associated with a reduced forced vital capacity (FVC, A). However, a significant correlation was not observed with diffusing capacity (B). Correlation analysis for FVC (A) revealed a r coefficient of −0.254, p = 0.026. For diffusing capacity (B), the r coefficient was −0.206, p = 0.074.

### Transplant-Free Survival

We then examined the association between three-year transplant-free survival and urinary desmosine concentrations. To create the comparator groups, we divided elastokine concentrations in patients with IPF into tertiles and assessed time to event (lung transplant or death) for 36 months from the first sample donation. We found that elevated desmosine concentrations were significantly associated with a worse clinical outcome (Figure 4, HR of 15.04, p=0.0005).

**Figure 4.**
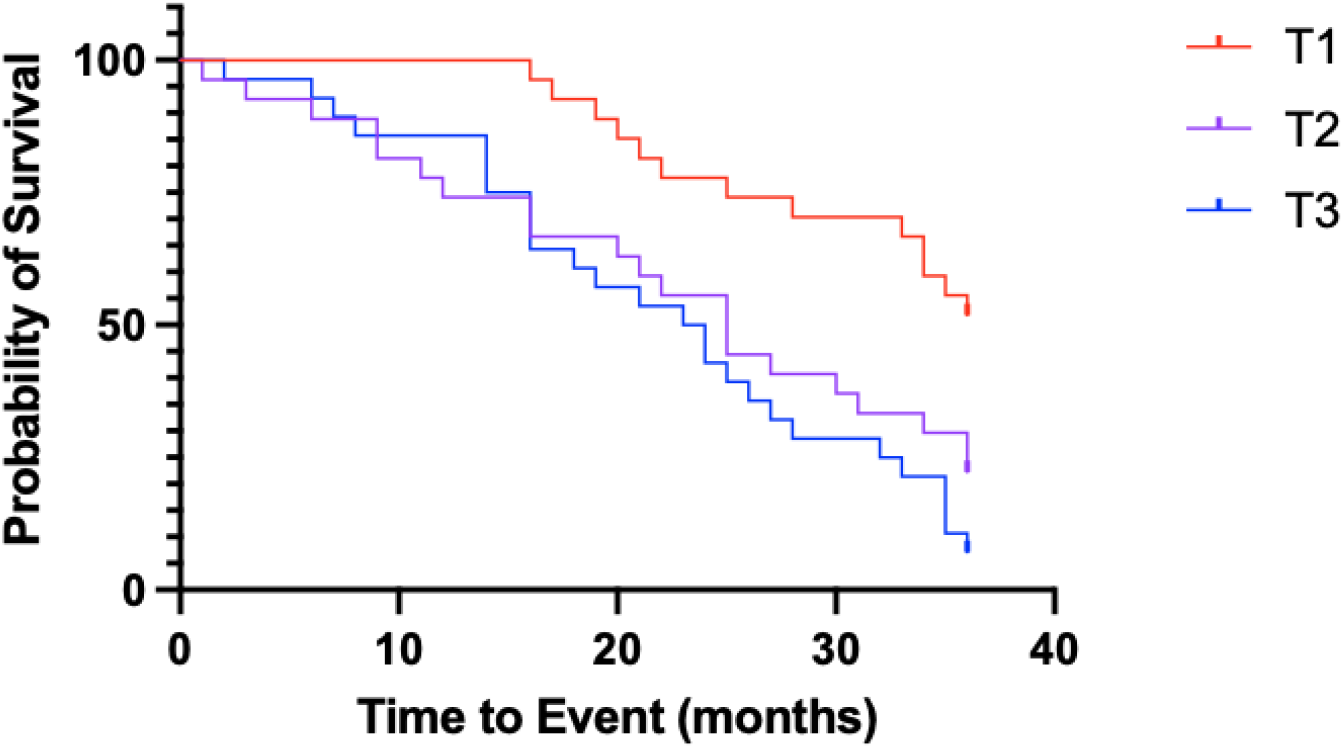
Three-year transplant-free survival. Higher concentrations of desmosine are associated with a worse transplant-free survival within 3 years of initial sample donation. Desmosing concentrations of patients with IPF were divided into tertiles and then Kaplan-Meier analysis was performed. HR (Mantel-Cox) 15.04, p = 0.0005.

### Elastokine Concentrations Associated with GAP Stage

The GAP (Gender, Age, and lung Physiology) staging system was initially created(42) and subsequently validated(43) to assess 1, 2, and 3-year mortality in patients with IPF. It uses a combination of gender, age, and two assessments of lung function (FVC and D_L_CO) to create a risk stratified stage. We assessed the association between GAP stage and elastokine elevation and found that the higher the elastokine level, the higher the GAP stage (Figure 5). This data further supports the association between elevated elastokines, IPF severity, and mortality.

**Figure 5.**
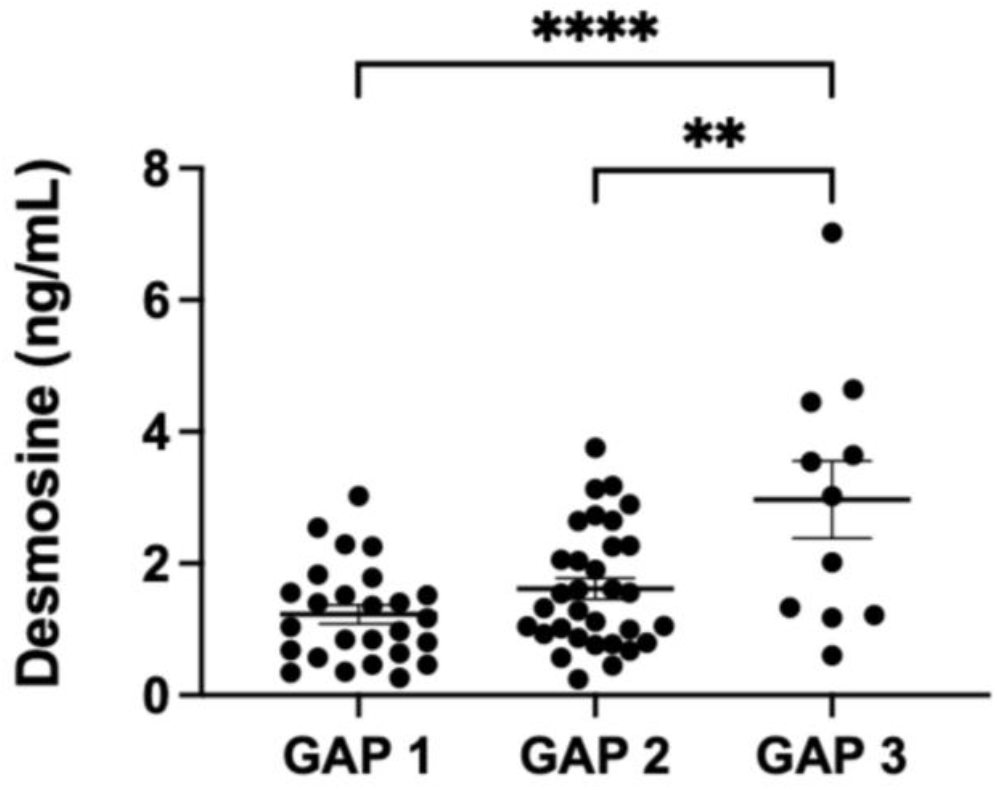
EDP concentrations and the association with GAP stage. Higher concentrations of desmosine are associated with a worse GAP stage. Comparison of GAP 2 to 3 p = 0.0018 and GAP 1 to 3 p < 0.0001.

## Discussion

Here we present data demonstrating an association between elastin remodeling, lung function, and disease severity in IPF. This work supports previous data demonstrating increased elastokine concentrations in serum and bronchoalveolar lavage fluid in patients IPF(45, 46), and our measurements are consistent with the level reported by de Brouwer et. al.(45). Our work highlights the association between higher concentrations of elastokine and clinical prognosis. We observed a significant association between elastokine elevation and decreased FVC, the primary index used to evaluate therapeutic efficacy in IPF. We also observed a similar, though not statistically significant, association between elastokine concentration and reduced D_L_CO. These associations make intuitive sense as one would anticipate decreased lung function due to replacement of elastin, which confers tissue resilience, deformability, and elasticity by comparatively stiffer collagen and other ECM proteins. We previously demonstrated a decreased ratio of mature elastin relative to collage in patients with IPF(47), which supports the hypothesis that mature elastin is replaced by collagen in pulmonary fibrosis. Perhaps more importantly, in this study, we also observed an association between elastokine elevation and reduced three-year transplant-free survival. A correlation between elastokine concentration and GAP stage, a surrogate for mortality, further supported this finding.

Our study also offers a potential practical advantage as the method of measurement we utilized is technically easier (ELISA assay), more widely available, and allows for high throughput processing compared to previously reported techniques that are more time, technical, and resource intensive(45, 46). We also demonstrate that desmosine and isodesmosine are readily detectable in both healthy volunteers and patients with IPF. In addition, there was good correlation between isodesmosine and desmosine concentrations regardless of the source (serum vs urine). As both were similarly elevated in patients in IPF, this suggests that elastin remodeling is the inciting event leading to release of these amino acids into the circulation. Finally, we also demonstrate that urinary and serum elastokine concentrations are consistently and similarly elevated in IPF. This adds to previous data measuring increased concentrations of circulating desmosine and isodesmosine in plasma(45) and bronchoalveolar lavage(46) samples in patients with IPF.

We acknowledge several limitations of the current study. First, we used a single-center retrospective cohort that comprised mostly Caucasian males. Although the sex distribution may be expected, we recognize that underrepresented minority patients are not included in our cohort and that our findings may not be generalizable to more ethnically diverse populations. Additionally, as the guideline-based definition of IPF evolved over the course of enrollment, some time-dependent discrepancies may have arisen in the clinical diagnosis of IPF. Some of our patients had combined pulmonary fibrosis and emphysema and/or WHO Group 3 pulmonary hypertension (<10% of our cohort), and it is unclear whether these co-morbid conditions contributed to the elevations in elastokine concentrations. We were unable to analyze associations between longitudinal changes in lung function, transplant-free survival, and elastokines due to an insufficient sample size. We were also unable to assess for longitudinal changes in elastokine concentrations as a function of anti-fibrotic medication for similar reasons. As expected, there was a significant difference in age between healthy volunteers and patients with IPF; however we did not have age-matched controls to differentiate between age-related and disease-related elastokine concentrations. Others have reported disease-specific increases above those expected from aging alone with respect to desmosine concentrations(22). Some degree of caution in interpreting our analyses is warranted due to the possibility of a Type I error given the relatively small sample size and the restricted geography from which biospecimens were obtained. However, our sample size was adequate based on our power calculation. In addition, our data is consistent with and builds upon previously published data. We plan to validate our findings with larger cohorts and, potentially, a prospective trial, ideally using comparative biomarker analysis.

There have been many promising biomarkers and clinical therapies, but for a variety of reasons, most do not achieve FDA approval(23). Emerging evidence suggests that elastokines are not simply a biomarker of lung tissue remodeling but might also contribute to lung pathology. In addition to desmosine, isodesmosine, other proteins are members of the elastokine family including α-elastin, κ-elastin, and a group of peptides with a conserved VGVAPG domain(24, 25). Elastokines have been shown to induce signal transduction and cell activation via several receptors including α_v_β_5_, galectrin-3, and integrins, but the majority of data suggests that the Elastin Receptor Complex (ERC) is probably the main receptor(25). This receptor is a heterotrimer composed of a peripheral subunit known as the Elastin Binding Protein, a protective protein/cathepsin A, and a membrane-associated protein called Neuraminidase-1. Elastokines have been shown to promote cell proliferation, invasion, survival, matrix metalloproteinase expression, angiogenesis, and fibroblast chemotaxis(26-32). Downstream signaling pathways of the ERC involve the MAP kinase family (ERK1/2) and pro-MMP-1 production(24). Preclinical administration of elastases is a well-established pre-clinical model of emphysema(33), however further research is needed into the mechanism(s) by which elastokines promote the development of emphysema and pulmonary fibrosis.

The progressive nature of IPF combined with the increasing world-wide incidence and prevalence of this disease(48), especially in the context of having only two partially effective therapies(3, 4), represents an unmet medical need. There are several potential novel therapeutics on the horizon, but many promising therapies have fallen short of demonstrating durable efficacy(23, 49). We propose that any additional insight into potential biomarkers and/or mechanistic understanding of this devastating disease will only help move the field of fibrosis research forward.

## Interpretation

Elevated concentrations of circulating elastokines, a marker of matrix turnover, are associated with worse lung function and reduced three-year transplant-free survival in patients with IPF. These results need to be confirmed with a validation cohort and studies examining the mechanism of EDP-induced fibrosis are currently underway.

## Data Availability

All data produced in the present study are stored on secure internal servers at URMC and are available upon reasonable request to the authors.

## Figures/Tables

**Table 1:** Baseline characteristics. Data comparing healthy volunteers and patients with IPF are presented as No. (%) with median and interquartile ranges for relevant parameters. Among those with IPF, patients were more likely to be male (71.6% vs 28.4%, p = 0.0001), Caucasian (97.5% vs 2.5%, p <0.0001), and have ever smoked (69.1% vs 30.9%, p = 0.0006).

| Variable | Healthy | IPF | p-value |
| --- | --- | --- | --- |
| **Age (Yrs)** | N=24 | N=81 | <0.001 |
| Median (IQR) | 40.0 (29.0 : 63.5) | 71.0 (66.0 : 79.0) |  |
| **Gender** | N=24 | N=81 | 0.004 |
| Male | 9 (37.5%) | 58 (71.6%) |  |
| Female | 15 (62.5%) | 23 (28.4%) |  |
| **Race** | N=24 | N=81 | 0.22 |
| White | 22 (91.7%) | 79 (97.5%) |  |
| Non-white | 2 (8.3%) | 2 (2.5%) |  |
| **Ever smoker** | N=24 | N=81 | 0.021 |
| No | 14 (58.3%) | 25 (30.9%) |  |
| Yes | 10 (41.7%) | 56 (69.1%) |  |
| **BMI** | N=22 | N=78 | 0.003 |
| Median (IQR) | 24.3 (23.3 : 28.0) | 28.0 (25.7 : 32.4) |  |
| **FVC (% predicted)** | N=23 | N=81 | <0.001 |
| Median (IQR) | 99.0 (91.0 : 108.0) | 72.0 (64.0 : 84.0) |  |
| **Diffusing capacity (% predicted)** | N=2 | N=80 | 0.08 |
| Median (IQR) | 81.5 (54.0 : 109.0) | 46.0 (37.0 : 54.5) |  |
| **Circulating Desmosine (ng/ml)** | N=22 | N=78 | <0.001 |
| Median (IQR) | 0.3 (0.2 : 0.4) | 1.4 (0.8 : 2.5) |  |
| **Oxygen needs** | N=24 | N=81 | <0.001 |
| No | 24 (100.0%) | 38 (46.9%) |  |
| Yes | 0 (0.0%) | 43 (53.1%) |  |
| **Anti-fibrotic** | N=0 | N=81 |  |
| No | 0 (0.0%) | 44(54.32%) |  |
| Yes | 0 (0.0%) | 37(45.68%) |  |
| **Death** | N=0 | N=81 |  |
| No | 0 (0.0%) | 28(34.57%) |  |
| Yes | 0 (0.0%) | 53(65.43%) |  |
| **Transplant** | N=0 | N=81 |  |
| No | 0 (0.0%) | 71(87.65%) |  |
| Yes | 0 (0.0%) | 10(12.35%) |  |

